# Scalp EEG reveals functional dissociable aperiodic timescales in divergence of mental health

**DOI:** 10.64898/2026.06.27.26355457

**Authors:** Yuhan Lu, Hangze Mao, Xing Tian

## Abstract

Understanding the divergence of mental health requires recognizing the brain’s real-time information processing. For example, aperiodic neural activity characterizing intrinsic brain dynamics is increasingly used as a biomarker in mental health, yet how its temporal complexity relates to mental deviations in ageing and disorders remains unknown. Here, using resting-state electroencephalography from approximately 1,700 participants across healthy, neurological and psychiatric disorders and chronic-pain cohorts, we show that the knee-like frequency structure recovered from individual spectra of neural activity converges into two reproducible, population-level temporal components. These slow and fast aperiodic timescales showed distinct functional profiles: the slow component remained stable across all states of mental health, whereas the fast component increased with healthy ageing, decreased in mental disorders and was unchanged in chronic pain. These findings establish that scalp EEG preserves functionally dissociable aperiodic timescales, possibly reflecting body-mind interactions, offering scalable, non-invasive temporal markers for quantifying divergent states of mental health.

## Introduction

The challenge in brain health is that the abnormality of cognitive functions does not always associate with apparent morphological changes in the brain^1^. Therefore, the key is to comprehend the brain operations that mediate normal cognitive functions as well as the impairments in these processes that give rise to disorders^2^. How the brain dynamically processes information is one of the most fundamental aspects that mediate normal cognitive functions^3,4^. The intrinsic neural dynamics can be quantified by the aperiodic component of the power spectral density (PSD) of neural activity (i.e., background 1/f-like decay as a function of frequency)^5,6^. Parameters of the aperiodic component, such as spectral exponents and knee frequency, have emerged as a powerful measure for indexing brain states, cognition, healthy ageing and neurological and psychiatric disorders (hereafter, mental-health disorders)^7–9^. However, the clinical application of aperiodic measures in brain health remains constrained by unclear theoretical guidance^9–11^.

Recently, a dual-timescale hypothesis has been proposed to explain the divergence of mental health from a perspective of body-mind interaction (**Fig. 1A**). The account hypothesizes that the brain operates across parallel temporal regimes: fast dynamics that support perception, cognition and action^12–15^, and slow dynamics that track bodily and physiological signals to maintain the stability of the internal body state^10,16^. Results in intracranial recordings support the account by showing that spectral knees resolve into two distinct aperiodic components^17–21^: a fast-timescale component aligned with cortical sensory-association gradients, and a slow-timescale component linked to physiological rhythms such as cardiac activity^17^. Connecting to the ‘allostasis-first’ conjecture that maintaining viable bodily states is a fundamental and stable brain function, whereas cognition-related operations are more vulnerable to biological and pathological constraints^22–24^, the dual-timescale hypothesis predicts that slow and fast neural dynamics would show differential sensitivity across various states of brain health.

**Figure 1.**
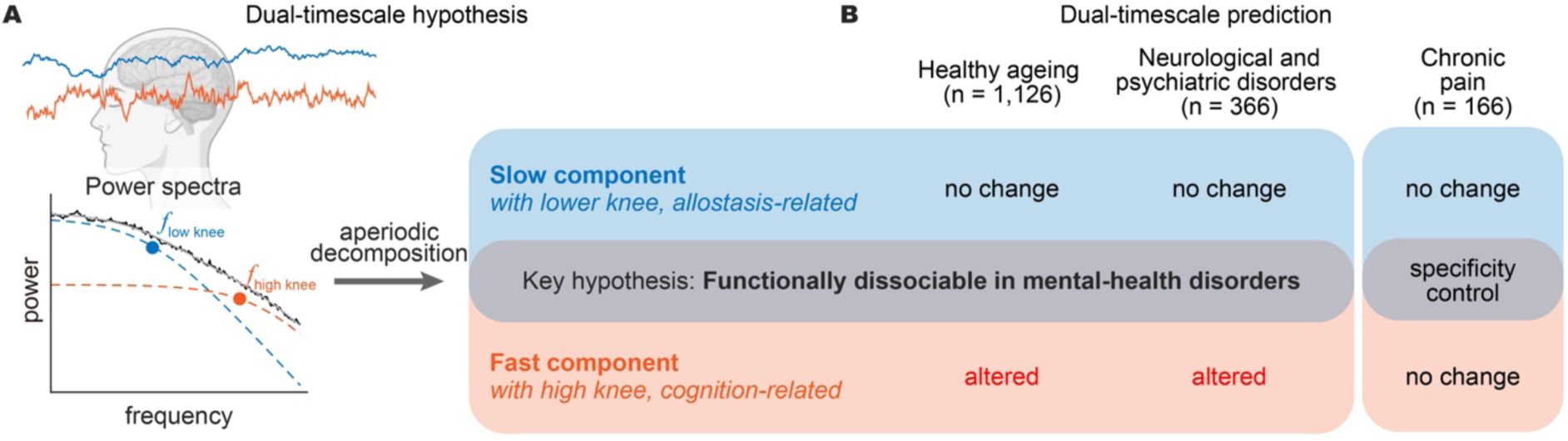
The dual-timescale hypothesis and predictions across states of brain health. **A**, The dual-timescale hypothesis states that intrinsic aperiodic neural activity comprises two parallel temporal components that mixes into a single power spectrum. Resting-state cortical activity (top) yields an aperiodic power spectral density (black) that is modelled as the sum of two Lorentzian components: a slow component with a lower-frequency spectral knee (*f*_low knee_, blue filled circle), and a fast component with a higher-frequency spectral knee (*f*_high knee_, orange filled circle). The two components can be recovered from the empirical spectrum by nested Lorentzian fitting (aperiodic decomposition, arrow). **B**, Predicted changes in the two components across states of brain health. According to previous studies, the slow component relates to the rhythms of internal bodily state, whereas the fast component relates to cognition. Following the ‘allostatic-first’ conjecture that maintaining viable bodily states is a more fundamental and comparatively protected brain function compared to the cognition-related dynamics, the two components are predicted to be functionally dissociable: the slow component is predicted to remain stable (no change) across all states of brain health, whereas the fast component is predicted to be state-sensitive (altered) in healthy ageing and mental-health disorders. Chronic pain, a clinically significant condition without a primary neurological or psychiatric diagnosis, serves as a specificity control. If the changes in the fast component reflects brain-disorder pathophysiology rather than a generic chronic-illness burden, the fast component would remain unchanged in the chronic pain cohort. n, number of participants per cohort.

Testing the dual-timescale account in the context of brain health requires non-invasive neurophysiological recordings in clinical populations. Scalp electroencephalography (EEG), a method that is widely used in cognitive, ageing and clinical settings, provides a feasible window. However, evidence for the dissociation of dual timescales has been obtained almost exclusively from intracranial recordings; whether scalp EEG preserves aperiodic dual-timescale components remains unknown. If dual timescales survive volume conduction and sensor-level mixing -- a critical assumption for non-invasive recording, they should show differential sensitivity across mental deviations in ageing and clinical conditions (**Fig. 1B**). Specifically, we predicted that the slow-timescale component would remain comparatively stable regardless of brain health status, whereas the fast-timescale component would be preferentially altered in healthy ageing and across various mental-health disorders.

Here, we combined a source-to-scalp generative model with nested Lorentzian fitting of resting-state scalp EEG from approximately 1,700 participants spanning healthy adults, brain disorder cohorts, and a chronic-pain comparison cohort. Across all cohorts, the recovered spectral knees consistently resolved into distinct slow- and fast-timescale components. These components showed the predicted functional dissociation: the slow component remained stable across all states of mental-health disorders, whereas the fast component was selectively altered by healthy ageing and the disorders. Notably, the fast component was preserved in a chronic-pain cohort that was included as a specificity control, suggesting that its alteration is specific to neurological and psychiatric pathophysiology rather than reflecting a generic effect of chronic-illness burden. Together, the functionally dissociable aperiodic timescales in scalp EEG index the body-mind interactions that impact mental operations, offering a novel temporal marker for non-invasively tracking the status of brain health.

## Results

### Multiple neural timescales are recoverable from scalp EEG as aperiodic knee frequencies

To establish whether multi-timescale neuronal dynamics could in principle survive volume conduction and produce a recognizable multi-knee signature at scalp EEG, we constructed a generative model in which 2,003 cortical sources tiling a template cortex were each driven by two independent Ornstein–Uhlenbeck currents. The summed unitary spectrum was, by construction, a sum of two Lorentzians with knees at 5 and 20 Hz (Fig. 2A). Sources were spatially correlated at biologically realistic levels^25^ and forward-modelled to 63 standard 10-10 electrodes through a three-layer BEM head model with realistic sensor noise; the resulting scalp signals were fitted with the nested multiple-Lorentzian fitting pipeline used on our experimental data (**Fig. S1**; see *Methods*). At the central Cz electrode the fit recovered two knees at 5.12 and 20.42 Hz, within 3% of the simulated 5 and 20 Hz (Fig. 2B, right), demonstrating that the underlying source timescales are quantitatively preserved at the scalp level. Across the full montage, 14 of 63 channels (22.2%) recovered the two-knee structure, with population median knees at 5.02 / 20.65 Hz; these channels clustered over central and parietal sites where summed source contributions to the lead-field were densest (Fig. 2B, left). Multiple distinct cortical timescales therefore remain detectable, and quantitatively faithful, on a substantial subset of scalp electrodes, supporting the interpretation that the multi-knee aperiodic structure observed empirically reflected multiple neural timescales rather than fitting artefact. We next estimated spectral knees in empirical resting-state EEG data and asked whether EEG knees separate into slow and fast population-level components.

**Figure 2.**
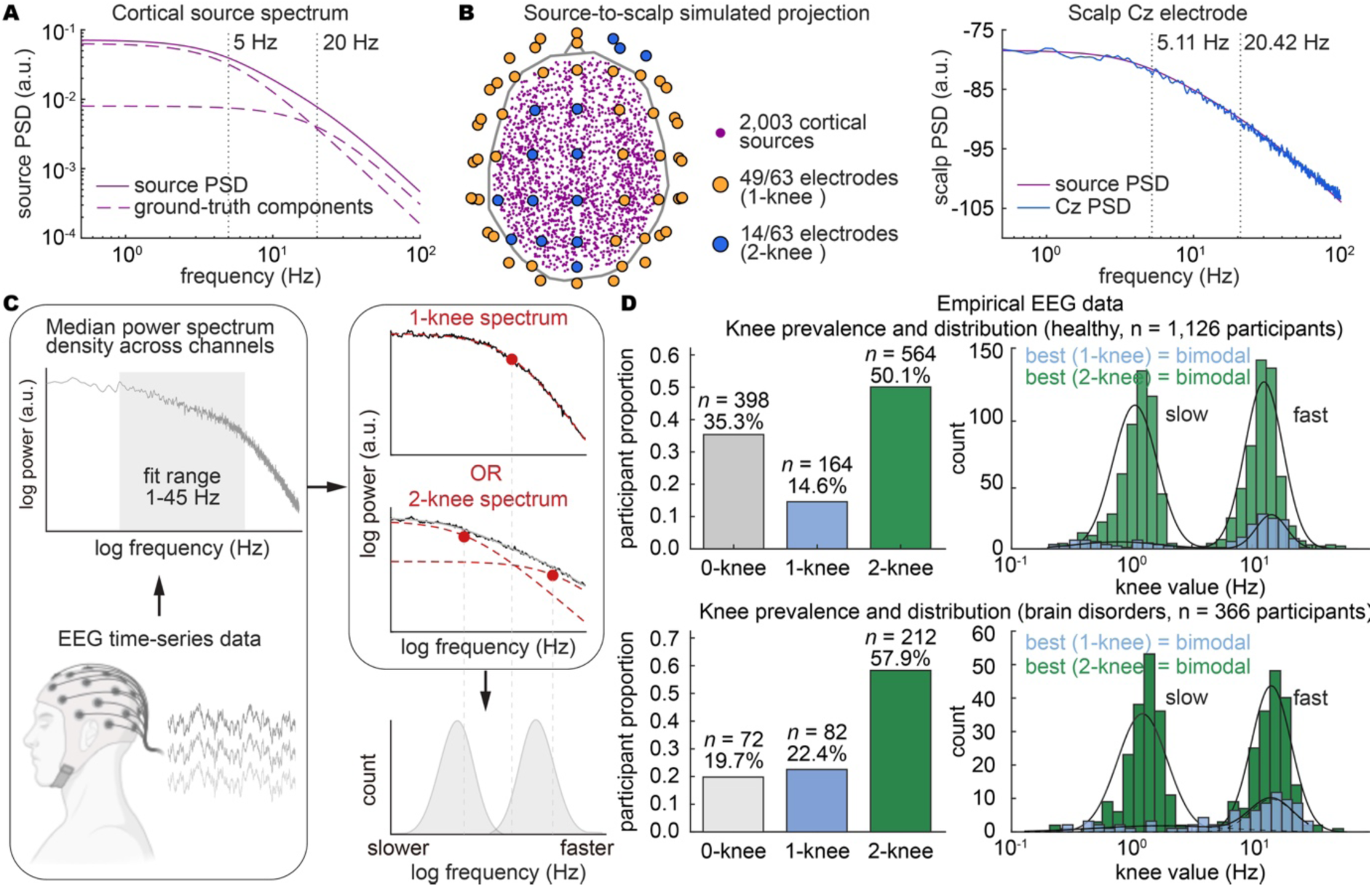
Testing theoretical and empirical multiple EEG aperiodic timescales. **A,** Per-source theoretical power spectrum. Two Lorentzian components with low and high knee frequencies (purple dashed lines) and their sum (purple line). Vertical dotted lines mark the simulated knee frequencies at 5 and 20 Hz, respectively. **B**, Simulation and results. Left, axial view of simulated 2,003 cortical source positions (purple), and 63 scalp electrodes colored by the number of recovered knees (orange, knee number = 1; blue, knee number = 2). Right, simulated PSD at the Cz electrode (blue); recovered knees 5.12 / 20.42 Hz, within 3% of ground truth. **C**, Procedures of empirical investigation in scalp EEG. Resting-state EEG spectra were first summarized as channel-median power spectra and fitted over 1-45 Hz using zero-, one-, or two-knee Lorentzian models. Each participant could therefore contribute either no knee, one knee, or two knees. All estimated knee frequencies were then pooled across participants to test whether they formed a single continuous distribution or two separable clusters. This population-level clustering provides evidence for whether EEG aperiodic activity is better described by one single continuous timescale or by two distinct components of timescales. The functional relevance of the slow- and fast-timescale components was further assessed by comparing their sensitivity to healthy ageing, mental-health disorders, and chronic pain. **D**, Empirical results of knee prevalence and population distribution. Across both healthy and brain-disorder cohorts, most participants showed at least one spectral knee, with two-knee spectra being the most prevalent class. Both pooled knee frequencies from one-knee and two-knee models were best explained by a bimodal distribution, revealing separable slow- and fast-timescale components at the population level.

### Empirical EEG knees resolve into slow and fast population components

Following the analysis procedural framework in Fig. 2C, we fitted each participant’s channel-median resting-state EEG spectrum with nested zero-, one- and two-knee Lorentzian models, allowing each participant to contribute zero, one or two empirical knee frequencies. Across 1,126 healthy participants, model selection identified no knee in 398 participants (35.3%), one knee in 164 participants (14.6%) and two knees in 564 participants (50.1%; Fig. 2D, upper), indicating that most participants showed at least one detectable knee and that two-knee spectra were the most common class. We then pooled the recovered knee frequencies across participants and tested whether they formed a single continuous distribution or separated into distinct clusters. In healthy participants, knees from one-knee spectra were best explained by a bimodal distribution, with a two-component bimodal distribution strongly favored over a one-component distribution (ΔBIC[unimodal-bimodal] = 190.28; Fig. 2D, upper). Pooling the low and high knees from two-knee spectra yielded the same structure (ΔBIC[unimodal-bimodal] = 1,028.02; Fig. 2D, upper). The two bimodal distributions showed almost identical intersections between two components, at 3.968 Hz for one-knee spectra and 3.972 Hz for two-knee spectra, indicating that knees recovered from different individual-level model classes converged onto the same two population-level frequency modes.

The same bimodal distribution was observed in mental-health disorders. Across 366 patients from six disorder cohorts, model selection identified one knee in 82 patients and two knees in 212 patients (Fig. 2D, lower), with two-knee spectra again forming the most prevalent class. Both the single-knee and pooled double-knee distributions were best explained by bimodal models (single-knee pool: ΔBIC[unimodal-bimodal] = 37.02; double-knee pool: ΔBIC[unimodal-bimodal] = 310.90; Fig. 2D, lower). The corresponding intersections between two components were 3.954 Hz and 3.861 Hz, closely matching the healthy-cohort intersections at 3.968 Hz and 3.972 Hz. Thus, empirical EEG knees consistently separated into slow and fast population-level components in both healthy and clinical cohorts (see also body-centered chronic pain in **Fig. S2**), providing evidence for two distinct aperiodic timescales in resting-state EEG.

Having established this population-level dual-timescale organization, we therefore treated knees according to their resolved timescale identity rather than the individual-level model class from which they were recovered. That is, slow- and fast-timescale knees were separated by a stable boundary near ∼3.9 Hz and were pooled across one-knee and two-knee spectra for subsequent analyses. We next asked whether these two components were functionally dissociable by testing whether they showed distinct sensitivity to healthy ageing, mental-health disorders and chronic pain.

### The slow-timescale component remains unchanged, but the fast-timescale component increases with healthy ageing

We next asked whether healthy ageing shifted both timescale components or selectively affects one of them (Fig. 3A). We first quantified potential influences of recording state and gender on component frequency. In paired recordings, eyes-open rest selectively increased the slow-timescale component relative to eyes-closed rest, with no detectable change in the fast-timescale component (*n* = 234; low EO/EC ratio = 1.077, *p* = 0.0227; high EO/EC ratio = 0.987, *p* = 0.64; Fig. 3B). In eyes-open recordings, men showed lower slow- and fast-timescale components than women after adjustment for age and dataset (n = 475; male/female ratio: low = 0.903, *p* = 0.00802; high = 0.881, *p* = 0.0001; Fig. 3B). Because these factors systematically influenced empirical knee estimates, subsequent ageing analyses were restricted to eyes-open recordings and included gender as a covariate.

**Figure 3.**
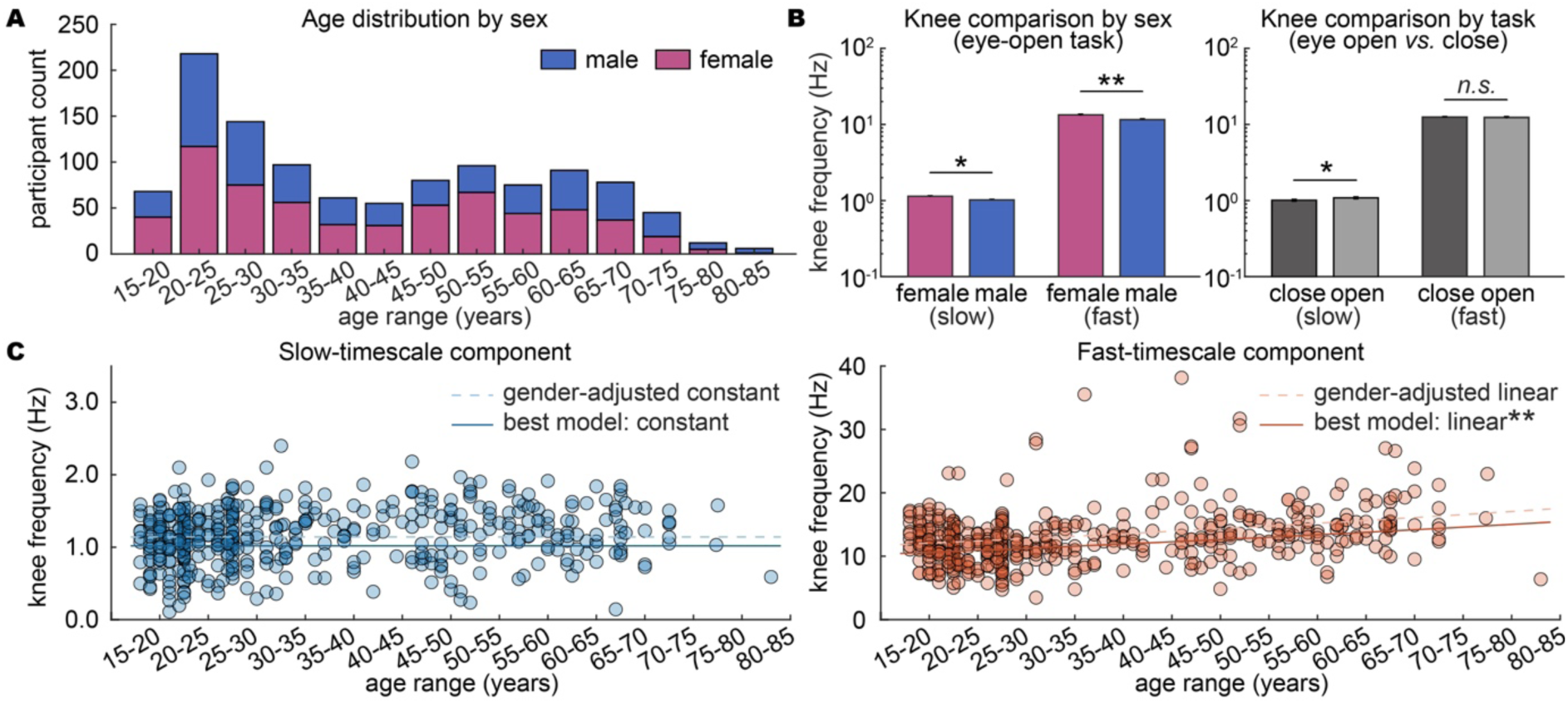
Healthy ageing selectively shifts the fast-timescale component. **A**, Age and gender distribution of the healthy cohort included in the component-wise analyses. **B**, Component frequencies stratified by gender and resting-state condition. Bars show group means; error bars denote s.e.m. **C**, Age associations for slow and fast components. Lines show fitted model trends. The slow component was best described by a constant model, whereas the fast component increased with age.

Ageing dissociated the two timescale components at the model-selection level. The slow-timescale component was best described by a gender-adjusted constant model, with no support for adding a linear age term (ΔBIC[linear−constant] = +1.33; Fig. 3C). By contrast, the fast-timescale component was best explained by a gender-adjusted linear model (ΔBIC[linear−constant] = −25.02; Fig. 3C), showing a robust increase with age (LR = 31.184, d.f. = 1, *p* = 2.35 × 10^-8^). This effect corresponded to a 9.8% increase per age standard deviation (16.2 years) and did not differ by gender (gender-by-age interaction: LR = 0.341, d.f. = 1, *p* = 0.559). The same dissociation was observed when one-knee and two-knee spectra were analyzed separately (**Fig. S3**), confirming that the effect did not depend on the individual-level model class from which knees were recovered. Thus, healthy ageing selectively increased the high-frequency, fast-timescale component while leaving the low-frequency, slow-timescale component comparatively stable.

### Mental-health disorders decrease the fast-timescale component, but the slow-timescale component remains unchanged

We next asked whether this fast-timescale component was also altered in mental-health disorders, against age- and gender-matched healthy controls within each cohort. The slow-timescale, low-frequency component showed no significant disorder effect in any types of brain disorder (Fig. 4A and 4C). By contrast, the fast-timescale component was consistently lower in patients than in matched controls across all six disorders (Fig. 4B and 4C). This reduction reached FDR-corrected significance in Alzheimer disease (disease/control ratio = 0.639, 95% CI = 0.531-0.769, *p* = 2.57 × 10^-4^, Cohen’s *d* = -1.24) and depressive disorder (ratio = 0.841, 95% CI = 0.753-0.940, *p* = 0.014, Cohen’s d = -0.42), with concordant but FDR-non-significant reductions in ADHD, insomnia, obsessive-compulsive disorder and Parkinson’s disease. (Fig. 4A**-C**). Thus, mental-health disorders selectively reduced the fast-timescale component, whereas the slow-timescale component remained stable. Together with the ageing results, the two timescale components are dissociated in states of mental health, and the fast-timescale component is the ageing and disorder-sensitive axis of the scalp EEG aperiodic dynamics.

**Figure 4.**
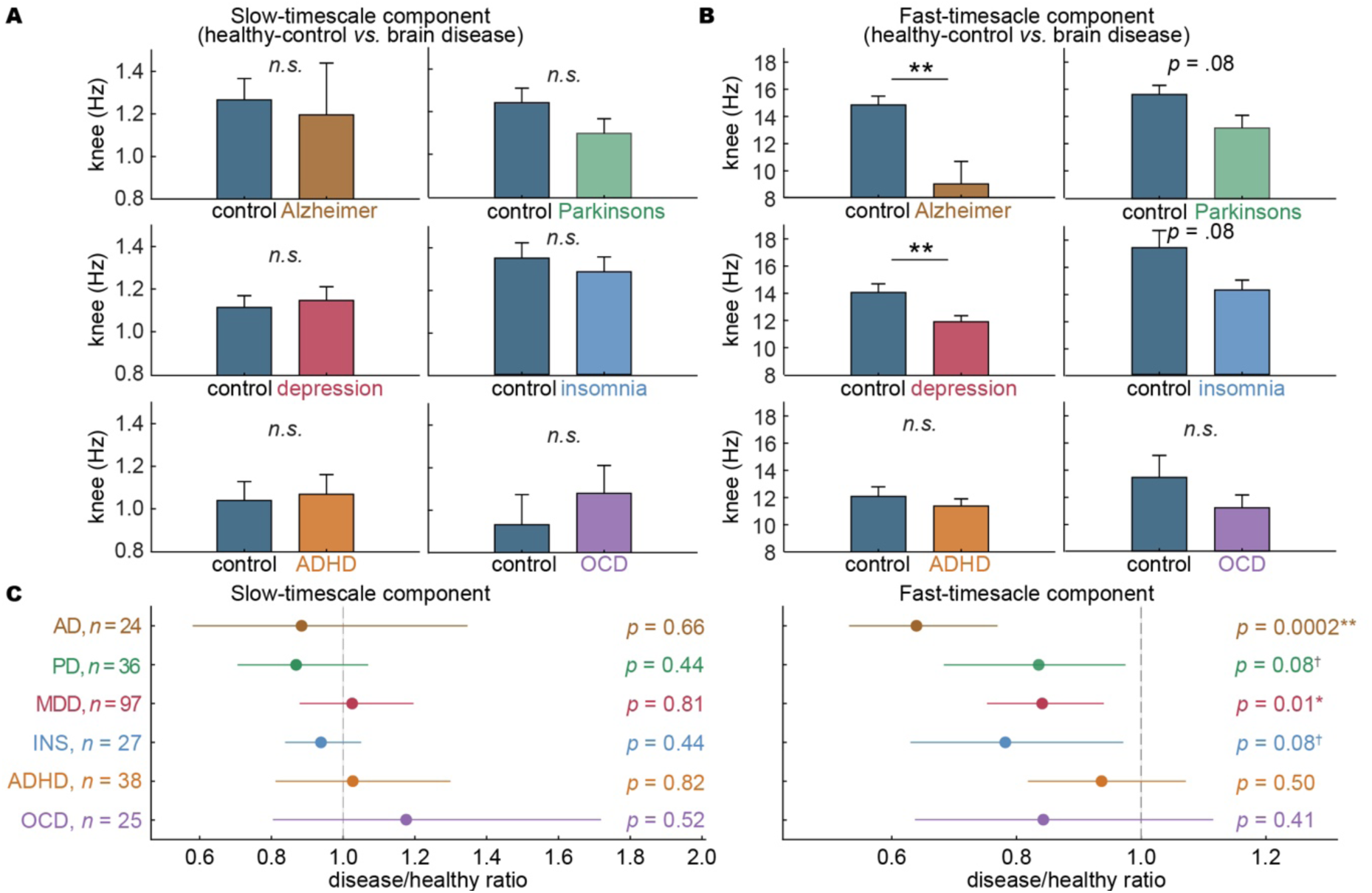
Mental-health disorders selectively reduce the fast component. **A**, Matched case-control comparisons of the slow-timescale component across six types of mental-health disorders. Bars show group estimates for patients and age- and gender-matched healthy controls; error bars denote s.e.m. **B**, Matched case-control comparisons of the fast-timescale component across the same six types of disorders. The fast-timescale component was lower in patients across disorders, with significant reductions in Alzheimer disease and depressive disorder after FDR correction. **C**, Forest plots summarizing disorder-to-control ratios for the slow- and fast-timescale components. Points show estimated ratios; horizontal lines denote s.e.m. Dashed vertical lines indicate no group difference. *P* values are FDR-corrected across disorders within each component. n.s., not significant; s.e.m., standard error of the mean; AD, Alzheimer’s disease; PD, Parkinson’s disease; MDD, major depressive disorder; INS, insomnia; OCD, obsessive–compulsive disorder; ADHD, attention-deficit/hyperactivity disorder; *p* < 0.05; *p* < 0.01.

### Chronic pain does not change fast component

Lastly, we asked whether any fast-component change reflects a generic effect of chronic-illness burden rather than neurological or psychiatric pathophysiology. To address this, we examined chronic pain as a body-centred comparison condition, because chronic pain represented a persistent and clinically significant disorder but was not primarily defined by a brain-disorder diagnosis. We compared chronic-pain participants with age- and gender-matched healthy controls. Neither component differed between groups: the slow-timescale component was unchanged in chronic pain (chronic pain/control ratio = 0.952, 95% CI = 0.825-1.098, *p* = 0.497), and the fast-timescale component was also unchanged (ratio = 1.014, 95% CI = 0.900-1.143, *p* = 0.815; Fig. 5A). This null pattern remained when chronic pain was divided into seven diagnostic subtypes (Fig. 5B): no subtype showed an FDR-corrected alteration in either component. Thus, chronic pain did not reproduce the fast-timescale component reduction observed in mental-health disorders, arguing against the fast-component being a generic marker of persistent bodily symptoms or chronic illness status. Instead, the results supported a more selective association between the fast-component alterations and brain disorder states. The slow-component also remained stable, reinforcing the dissociation between the two components: the fast component was selectively altered in both healthy ageing and mental-health disorders, whereas the slow component was comparatively preserved across healthy ageing, mental-health disorders and chronic pain.

**Figure 5.**
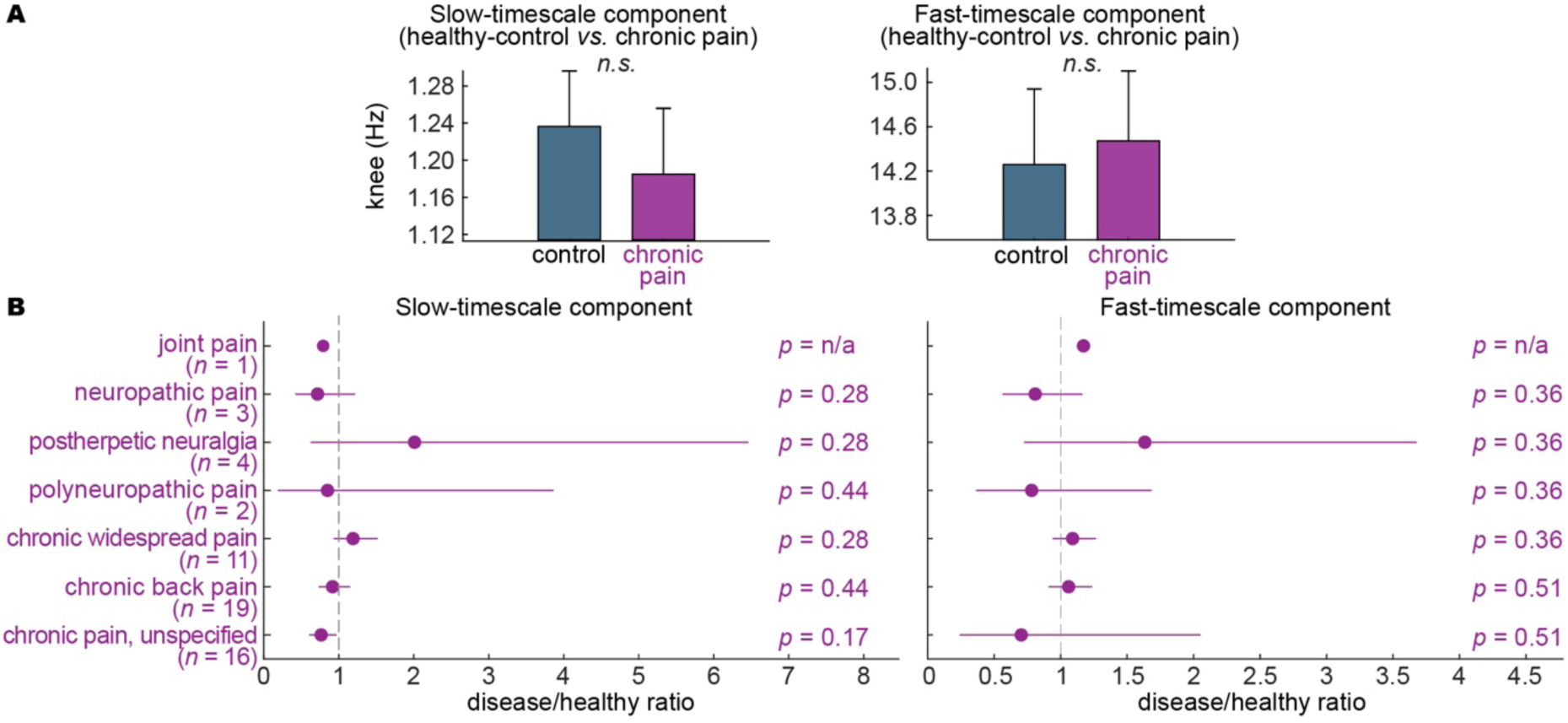
Chronic pain retains the two-timescale components without altering either component. **A**, Matched case-control comparisons of slow and fast components between chronic-pain participants and age- and gender-matched healthy controls. Bars show group estimates; error bars denote s.e.m.; Neither component differed between groups. **B**, Forest plots summarizing chronic-pain subtype-to-control ratios for slow and fast components. Points show estimated ratios; horizontal lines denote s.e.m.; Dashed vertical lines indicate no group difference. P values are FDR-corrected across chronic-pain subtypes within each component. n.s., not significant.

## Discussion

A central challenge in human neural dynamic research is that the physiological meaning of scalp-recorded aperiodic activity remains unclear, especially in clinical settings. Although spectral exponents and knees are increasingly used as biomarkers of ageing, cognition and mental disorders, most studies implicitly treat the aperiodic spectrum as a single process that can be summarized by one exponent or one characteristic timescale. Here, EEG spectral knees consistently resolved into two population-level timescale components across healthy adults and clinical cohorts: a slow component (<∼4 Hz) and a fast component (>∼4 Hz). While the slow component remained largely stable across ageing and mental-health disorders, the fast component systematically differentiated the ageing from the disorders. This dissociation reveals previously unrecognized structure in scalp EEG aperiodic activity.

Rather than reflecting a purely methodological limitation, our findings suggest that a major source of ambiguity in the aperiodic EEG literature may stem from how the signal itself is conceptualized. Most studies interpret the aperiodic spectrum as the expression of a single underlying process and therefore summarize it using one exponent or one knee^5,6^. Under this framework, observed differences are typically attributed to changes in the magnitude of that process. However, the present results indicate that scalp EEG contains at least two separable aperiodic timescales with distinct functional characteristics^17–21^. If these components are combined into a single estimate, changes in one component may be masked, diluted or distorted by the stability of the other^17^. This provides a plausible explanation for why aperiodic markers have often shown heterogeneous effects across ageing and clinical populations despite being proposed as general indicators of brain health^10,16,26^. Extended from previous intracranial recordings showing separable slow and fast aperiodic components, our source-to-scalp simulations and empirical results demonstrate that these components survive the spatial mixing, skull filtering and volume conduction that shape scalp EEG.

The key advance is not only that two aperiodic components can be detected in scalp EEG, but that they show distinct functional profiles. This dissociation is consistent with our previous intracranial study^17^, in which sEEG recordings showed that fast aperiodic timescales followed cortical functional gradients, whereas slow aperiodic components were associated with heart-related cortical potentials and physiological rhythms. The present EEG study supports the slow-fast distinction from a different angle: fast timescales changed with healthy ageing and mental-health disorders, whereas slow timescales remained comparatively stable across healthy ageing, mental-health disorders and chronic pain. This pattern is consistent with allostatic theories of brain-body regulation, in which maintaining viable bodily states is a fundamental and protected function, whereas cognition-related operations are more vulnerable to biological and pathological constraints^22–24^. Under this view, the slow component may mediate a stable allostasis-related regulatory function, whereas the fast component may reflect a more flexible cognition-related axis that changes with ageing and brain-related dysfunction. The distinct evolving profiles in both temporal components are manifested in different states of mental health.

The fast component was sensitive to both healthy ageing and mental-health disorders, but in opposite directions -- increasing with healthy ageing and decreasing in mental-health disorders. The divergence suggests that healthy ageing and brain-related pathology should not be treated as equivalent shifts along a single nonspecific deterioration axis. Instead, healthy ageing may reflect a gradual acceleration of cortical temporal dynamics, consistent with shorter autocorrelation, increased neural variability or excitation-inhibition rebalancing^27–29^, whereas mental-health disorders may disrupt the fast-timescale organization that supports cognition-related cortical function^9^. Thus, the fast component provides a scalable, non-invasive temporal marker that separates normative ageing-related change from pathological brain dysfunction.

Together, our findings reframe scalp EEG aperiodic activity from a single-index biomarker into a functionally dissociated dual-timescale architecture. The distinct evolving profiles in slow and fast temporal components highlight the possible body-mind interaction in the brain dynamics that relates to mental health. Decomposing scalp aperiodic activity into slow and fast components offers a mechanistic framework for improving the specificity of EEG-based biomarkers in quantifying divergent states of brain health.

## Methods

### Study design and datasets

Resting-state EEG from ∼1,700 participants across healthy, mental-health-disorder and chronic-pain cohorts was analyzed with a harmonized pipeline comprising EEG preprocessing, IRASA-based spectral separation, nested Lorentzian knee fitting, construction of slow and fast knee components, and statistical modelling. The knee-estimation procedure was validated in two simulations: a source-to-scalp generative model testing whether cortical knees survive volume conduction and remain recoverable at the scalp (see *Simulation: source-to-scalp generative model*), and a synthetic-spectrum benchmark quantifying its accuracy against spectra with known ground truth (see *Simulation: fitter validation on synthetic spectra*).

Public EEG data for the primary analyses were obtained from nine openly available resources: seven BIDS-formatted OpenNeuro datasets, including ds002778, ds003490, ds003775, ds004148, ds004504, ds004902 and ds005385^30–37^; the Leipzig Mind-Brain-Body (LEMON) dataset^38^; and the TDBRAIN database^39^. Disease analyses included 366 participants from public EEG cohorts labelled as Parkinson’s disease, Alzheimer’s disease, ADHD, insomnia, depressive disorder or obsessive-compulsive disorder. Parkinson’s disease data were obtained from OSF_pehj9^40^, OpenNeuro ds002778^31^, OpenNeuro ds003490 and TDBRAIN; Alzheimer’s disease data were obtained from OpenNeuro ds004504; and ADHD, insomnia, depressive disorder and obsessive-compulsive disorder data were obtained from the corresponding TDBRAIN diagnostic labels. Chronic-pain analyses were included as a specificity control and comprised 166 chronic-pain participants and 81 healthy controls from OSF_m45j2 and OSF_srpbg^41,42^. In analyses requiring matched healthy controls, the analytic sample size was determined by the availability of suitable age- and gender-matched controls. Therefore, the final number of included participants could be smaller than the total cohort size when no eligible matched control was available for a given participant.

### EEG data preprocessing

All processing was performed using custom MATLAB scripts adapted from established analysis pipelines^43,44^. Preprocessing was performed at the channel-by-time level before any participant-level spectral aggregation. Signals sampled at ≥500 Hz were resampled to 500 Hz; datasets sampled below 500 Hz were retained at their native sampling rate and analyzed within the corresponding Nyquist-limited frequency range. Line noise was attenuated before bad-channel detection and ICA using narrow-band notch filters at the dataset-appropriate line frequency, typically 50 Hz, and its harmonics up to 200 Hz. Notched frequencies were excluded from subsequent spectral model fitting. Bad channels were detected using robust outlier criteria based on amplitude dispersion, inter-channel correlation and residual line noise. Specifically, channels were rejected for abnormal log standard deviation or log median absolute deviation, low median correlation with other scalp channels, or excessive residual line noise after notch filtering. Correlation-based rejection was capped at 35% of scalp channels to avoid excessive rejection in datasets with globally low inter-channel correlations. Average referencing was then performed using retained good channels, with iterative bad-channel screening and a minimum of eight channels required for reference estimation. ICA-based artefact attenuation was applied to good channels when at least 12 retained scalp channels were available. ICA was trained on data high-pass filtered at 1 Hz, down-sampled to 250 Hz when applicable, and capped at 150,000 training samples. The number of components was limited by data rank, channel count and a maximum of 32 components. Candidate ocular, muscle and cardiac components were automatically scored using spatial, spectral and temporal features, with EOG-proxy criteria used when available. To prevent overcorrection, removal was capped at two ocular, three muscle and one cardiac component. ICA-cleaned data were retained only when the relative reconstruction RMS error was ≤0.10; otherwise, the re-referenced signal before ICA was used for downstream analyses.

### Computing the aperiodic spectrum

The aperiodic spectrum was isolated using Irregular-Resampling Auto-Spectral Analysis (IRASA)^45^, which separates the fractal (aperiodic) component of a power spectrum from superimposed oscillations by exploiting their different behavior under resampling. IRASA was implemented with the amri_sig_fractal function applied to each retained scalp channel. Preprocessed data were segmented into 30-s windows with 50% overlap. Windows were screened for artefacts using robust outlier criteria on window-level RMS and peak amplitude, and were retained when both robust z-scores were ≤3.5; if this removed more than half the windows, the least extreme windows were restored to retain at least 50% of windows, or at least eight windows when available. For each retained channel and window, IRASA was applied over 0.1–100 Hz with detrending and anti-alias filtering enabled and default resampling factors from 1.1 to 1.9 in steps of 0.05, decomposing the spectrum into mixed, fractal and oscillatory components; the fractal component was taken as the aperiodic spectrum. Channel- and window-level fractal spectra were then aggregated to a single spectrum per participant and condition. We first defined a core set of scalp channels spanning frontal, central, temporal, parietal and occipital regions; the median fractal spectrum was computed across these core channels when enough were retained, and otherwise across all retained good scalp channels. The resulting participant-condition median fractal spectrum (median across retained windows and selected channels) was the input to nested Lorentzian fitting.

### Nested Lorentzian fitting

#### Algorithmic design

We estimated spectral knees by fitting power spectra with nested generalized Lorentzian models allowing zero, one or two knees. For empirical data the input was the IRASA fractal spectrum (above); for simulations, the Welch PSD (Simulation: Source-to-scalp generative model) as no oscillatory components were mixed into OU signals. Fits were performed in the 1-45 Hz band, while candidate knees were constrained to the broader search range 0.1-100 Hz. The exponent was fitted from the data and, for two-knee models, was shared across low- and high-knee components.

The zero-knee model was a power law in log-log space:

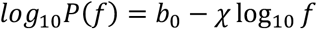

The one-knee model was:

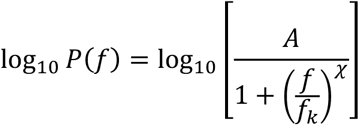

The two-knee model was:

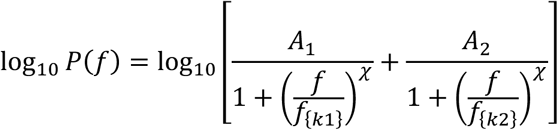

with fk1 < fk2 and a shared fitted exponent chi. Fits minimized residual error in log10 PSD. When a spectrum was summarized from multiple windows, the median across windows in log-PSD space was used and the window-to-window log-PSD variance was retained as a diagnostic weight; spectra were then restricted to the fitting band and interpolated onto a uniform log-frequency grid.

The one-knee model used 64 log-spaced knee seeds across the search range and exponent seeds [0.75, 1, 1.5, 2, 3, 4]. The two-knee model used a 10 × 10 ordered knee grid, a seed budget of the 20 best two-knee seeds, the same exponent seed pool, and a minimum log10 knee separation of 0.01. Parameters were optimized with fminsearch using tight tolerances. Model selection was nested: the one-knee model was first compared against the zero-knee model, and the two-knee model was then compared against the selected one-knee model.

The selection score was a BIC-type score, *BIC* = *nlog*(*MSE*) + *klog*(*n*), where n is the number of fitted log-frequency samples and k is the number of fitted parameters. The zero-to-one and one-to-two BIC margins were both 0 in the formal run.

To reduce low-frequency boundary-driven two-knee calls, a secondary stability criterion was enabled. If a selected two-knee fit contained a weak component whose integrated fractional contribution was < 0.15, the fit was downgraded to the one-knee model. If the selected two-knee fit had a low knee below 2 times the lower fit frequency, a conditional window bootstrap was run when multiple PSD windows were available. Twelve bootstrap resamples were fitted, and the two-knee model was retained only if at least 67% of bootstrap fits also selected two knees. Output files stored selected model order, knee frequencies, fitted exponent, fitted PSD, BIC-type scores, and R^2^.

### Construction of slow and fast knee components

Participant-level analyses used one representative record per participant. To define population-level temporal components, the recovered knee frequencies were pooled across participants and their distribution was characterized in log10 Hz: one-knee participants contributed their single finite positive knee, and two-knee participants contributed both their low and high knees. Pooled knee frequencies were fitted with Gaussian mixture models of one, two or three components and compared by BIC; a multi-component solution was reduced to a unimodal label when any component had low weight (<0.10) or when component separation was below one pooled standard deviation. Both the one-knee and pooled two-knee distributions across cohorts were best described as bimodal, and the intersection between the two fitted Gaussian components defined the slow/fast cutoff.

### Statistical analysis

#### General statistical framework

All group-level analyses were performed on log-transformed knee frequencies. Regression coefficients were back-transformed to frequency ratios on the Hz scale. Unless otherwise stated, statistical inference used two-sided Wald tests for the coefficient of interest, and 95% confidence intervals were obtained from model coefficient intervals and back-transformed for ratio reporting. Linear mixed-effects models were fitted using maximum likelihood. When the planned random-effects structure was not supported by the data or failed to converge, progressively simpler random-effects structures were used, followed by linear models when necessary. Multiple comparisons were controlled using Benjamini-Hochberg false-discovery-rate correction within prespecified families of tests.

#### Eyes-open versus eyes-closed comparisons

Condition comparisons included only participants with both eyes-open and eyes-closed recordings and valid two-knee fits in both conditions, with neither knee located at the search boundary. Low and high knees were analyzed separately using the model: *log*_10(*knee frequency*) ∼ *condition* + (1 | *participant*) + (1 | *dataset*). Eyes closed was used as the reference condition, and the eyes-open coefficient was the effect of interest. The formal healthy analysis included 234 paired participants. Effects were reported as eyes-open/eyes-closed frequency ratios with 95% confidence intervals.

#### Gender comparisons

Gender comparisons were performed in eyes-open records only and were restricted to one-and two-knee fits in which no knee was located at the search boundary (i.e. not at the 0.1–100 Hz boundary), with available age and gender information. Low and high knees were modelled separately using: *log*_10(*knee frequency*) ∼ *gender* + *age*_*z* + *age*_*z*^2 + (1 | *dataset*). Female was used as the reference group, and the male coefficient was the effect of interest. The formal analysis included 269 female and 206 male participants.

#### Age associations

Primary age analyses were performed on eyes-open participant-condition records. Records were required to have finite age information and binary gender labels. This yielded 475 included eyes-open records from 970 available eyes-open records. Exact age was used when available. For datasets reporting age ranges, the midpoint of the age bin was used for modelling, and a 5-year age bin was retained for visualization. Low- and high-knee age associations were modelled separately using mixed-effects models on log10 knee frequency. All candidate models included gender as a fixed covariate and dataset as a random intercept. The age effect was selected from a prespecified model family containing no age term, linear age, quadratic age and cubic age, with age z-scored before modelling. Models were ranked by BIC, and the simpler model was selected when candidate models differed by less than 2 BIC units. When the selected model included an age term, a gender-by-age interaction of the same polynomial order was tested using likelihood-ratio comparison. Parallel pooled-component age analyses used the healthy-cohort slow/fast component labels and age-bin midpoints. The same candidate model family was fitted separately for knees originating from one-knee and two-knee records.

#### Comparisons in disorders

Analyses of disorder cohorts compared each diagnostic group with condition-matched healthy controls. Eyes-open records were used for all diagnostic groups except Alzheimer disease (and its matched healthy control), for which eyes-closed records were used because eyes-open recordings were unavailable. Eligible records included either one-knee records with a finite positive knee or two-knee records with finite positive low and high knees.

Healthy controls were matched to participants in disorders at the participant level without replacement using exact gender matching and a 5-year age caliper. Controls were selected from healthy datasets and prioritized by smallest age difference, followed by higher model R^2^. The matched samples were: ADHD, 38 disease and 38 control participants; Alzheimer disease, 24 and 24; depressive disorder, 97 and 97; insomnia, 27 and 27; obsessive-compulsive disorder, 25 and 25; and Parkinson’s disease, 36 and 36. After matching, knees were assigned to slow or fast components using the healthy-cohort mixture cutoffs. Group differences were then tested separately within each component on log10 knee frequency. The preferred model was: *log*_10(*knee frequency*) ∼ *group* + *age*_*z* + *gender* + (1 | *dataset*) + (1 | *matc*ℎ*ed pair*). The disorder-group coefficient was the primary effect of interest. Benjamini–Hochberg FDR correction was applied across disorder-by-component tests.

#### Analysis in chronic-pain cohort

Analyses in chronic-pain cohort used eyes-open records only. The primary chronic-pain comparison was restricted to two-knee records with valid age, gender and fit information. Healthy controls were matched 1:1 without replacement using exact gender matching and a 5-year age caliper. Matching was performed in two stages. Internal healthy controls from the chronic-pain dataset were used first, and unmatched chronic-pain participants were then matched to legacy external healthy eyes-open controls. The final matched sample included 56 chronic-pain and 56 healthy participants, comprising 32 internal and 24 legacy matched pairs. Low and high knees were analyzed using the same mixed-effects structure as the disorder analyses, with chronic pain as the group coefficient of interest. These analyses were treated as specificity-control comparisons and were reported with corrected p values.

### Simulation: Source-to-scalp generative model

#### Source-level dynamics

To generate aperiodic activity with known ground-truth knees, we modelled each cortical source as a sum of independent Ornstein–Uhlenbeck (OU) processes. A single OU process had a Lorentzian power spectrum with one spectral knee set by its timescale, so a sum of OU components yielded a spectrum that was, by construction, a sum of Lorentzians with knees at prescribed frequencies. We simulated source-level cortical activity on 2,003 vertices of a decimated fs_LR cortical template^46^. Each source was driven by two independent zero-mean OU components, with knee frequencies of 5 and 20 Hz. The corresponding timescales were defined as τ = 1/(2πf)^47^. OU processes were sampled at 1,000 Hz using the exact discrete-time solution with a 1-ms integration step.

Spatial correlations were introduced by mixing source-specific OU fluctuations with spatially smooth shared OU modes. For each timescale, 16 independent global modes were generated, and their contribution to each cortical source decayed with distance according to a Gaussian kernel with a 60-mm coherence width. The shared-mode weight was set to ρ = 0.12, yielding a mean pairwise source correlation of 0.103, within the biologically plausible range reported for cortical dipoles^25^. Component amplitudes were set to [1, 0.5]. Each simulated dataset lasted 300 s.

#### Forward model

A three-layer boundary-element head model (FieldTrip standard_bem^48^; MNI template) and 63 standard 10-10 electrode positions were used. The lead-field matrix L (63 × 2,003) was computed with ft_prepare_leadfield and projected onto the per-vertex outward cortical normal so each dipole was orthogonal to the cortical surface. Scalp signals were X = LS + N with channel-independent Gaussian noise N at 26 dB variance-SNR per channel.

#### Spectral estimation and multi-knee fitting

Per-channel spectra were estimated by Welch’s method (5-s Hamming windows, 50% overlap) and fitted over 1–45 Hz with the nested Lorentzian procedure described above (see *Nested Lorentzian fitting*), without parameter tuning.

### Simulation: Fitter validation on synthetic spectra

We validated the nested Lorentzian fitting procedure using simulations with known zero-, one- and two-knee spectra. Synthetic spectra were generated from OU processes so that the ground-truth number and location of spectral knees were known. The simulation benchmark used the same fitting settings as the EEG analysis pipeline, including the 1-45 Hz fitting band, 0.1-100 Hz knee search range, multi-start exponent and knee initialization, nested BIC-based model selection, weak-component rejection and low-frequency boundary stability checks.

The benchmark comprised 38,932 ground-truth spectral conditions, with 10 independent repetitions per condition, yielding 389,320 fitted simulations. Single-knee spectra were generated with ground-truth knees from 0.2 to 60 Hz in 0.2-Hz steps. Two-knee spectra combined low knees from 0.2 to 10 Hz in 0.05-Hz steps with high knees from 12 to 90 Hz in 0.4-Hz steps. Each simulated time series was 300 s long, divided into 30-s windows, and fitted using the same window-wise aggregation procedure as the empirical EEG data.

Simulation performance was quantified by model-order accuracy, zero-, one- and two-knee selection rates, single-knee mean absolute error, and low- and high-knee estimation errors for correctly identified two-knee spectra. These simulations were used to characterize the parameter regimes in which the fitter reliably recovered the correct model order and knee frequencies, and to identify expected failure modes, including close knees, boundary-adjacent knees and knees outside the observable fitting band. These results motivated conservative handling of boundary-hit fits in the empirical analysis.

## Data Availability

Public EEG data for the primary analyses were obtained from nine openly available resources: seven BIDS-formatted OpenNeuro datasets, including ds002778, ds003490, ds003775, ds004148, ds004504, ds004902 and ds005385; the Leipzig Mind-Brain-Body (LEMON) dataset; and the TDBRAIN database.

https://openneuro.org/datasets/ds002778

https://openneuro.org/datasets/ds003490

https://openneuro.org/datasets/ds003775

https://openneuro.org/datasets/ds004148

https://openneuro.org/datasets/ds004504

https://openneuro.org/datasets/ds004902

https://openneuro.org/datasets/ds005385

https://fcon_1000.projects.nitrc.org/indi/retro/MPI_LEMON.html

https://brainclinics.com/resources/

## Data and materials availability

All data analyzed in this manuscript are from open data sources, subject to the access terms of the original repositories. The MATLAB codes and plots supporting the findings of this study are available at https://github.com/.

