## Supplementary documents for "Scalp EEG reveals functional dissociable aperiodic timescales in divergence of mental health"

### Supplementary Document

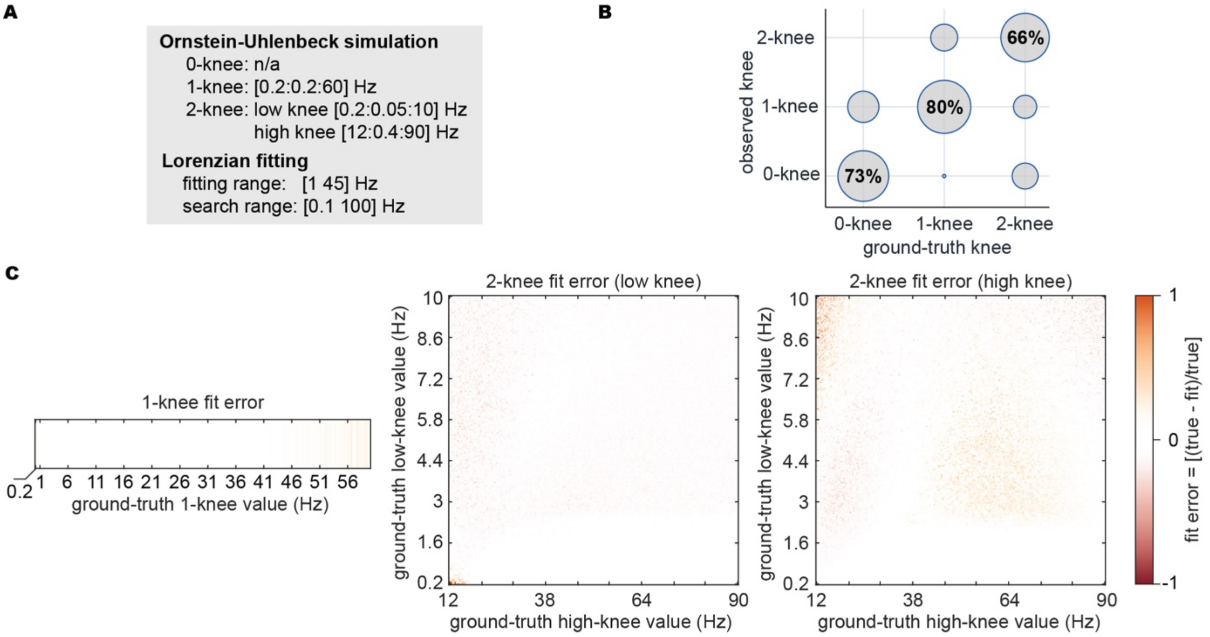


**Supplementary Figure 1.** Simulation validation of nested Lorentzian fitting for recovering aperiodic knees from a limited EEG fitting band. **A**, Simulation and fitting settings. Synthetic Ornstein–Uhlenbeck spectra were generated with zero, one or two ground-truth spectral knees. Single-knee spectra contained knees from 0.2 to 60 Hz, whereas two-knee spectra combined low knees from 0.2 to 10 Hz with high knees from 12 to 90 Hz. All spectra were fitted using the same nested Lorentzian pipeline as the empirical EEG analysis: a 1-45 Hz fitting band, a broader 0.1-100 Hz knee search range, and zero-, one- and two-knee candidate models. **B**, Model-order recovery. Bubble positions indicate the selected model order relative to the ground-truth model order; bubble size indicates the proportion of simulations assigned to each cell. Percentages on the diagonal show correct recovery rates for zero-, one- and two-knee spectra. **C**, Knee-frequency estimation error. The left panel shows relative fitting error for single-knee spectra, and the middle and right panels show relative errors for the low and high knees in two-knee spectra. Error was defined as ($knee_{true} – knee_{fit}$) / $knee_{true}$. Knees within the 1-45 Hz fitting band were recovered most accurately, whereas knees outside the fitted band showed larger errors but remained partially recoverable from the curvature they imposed within the observed spectrum. These simulations define the parameter regimes in which the nested Lorentzian procedure can recover broad aperiodic knee structure from scalp EEG spectra.

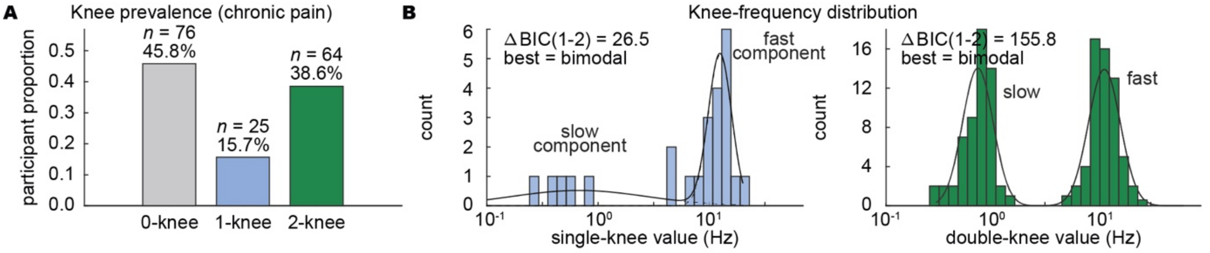


**Supplementary Figure 2.** Chronic pain preserves the two-component aperiodic knee structure. **A**, Prevalence of selected model order in chronic-pain resting-state EEG. Bars show the proportion of chronic-pain participants whose spectra were best described by zero-, one- or two-knee Lorentzian models. A substantial fraction of participants showed at least one detectable knee, and two-knee spectra were common. **B**, Population-level knee-frequency distributions in chronic pain. Recovered knees from one-knee spectra and pooled knees from two-knee spectra were both better explained by bimodal than unimodal distributions, revealing separable slow- and fast-timescale components.

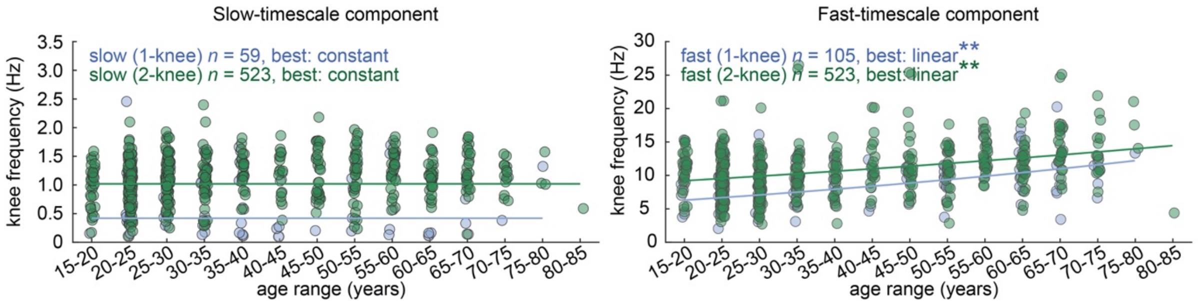


**Supplementary Figure 3.** The ageing dissociation between slow- and fast-timescale components is preserved when one-knee and two-knee spectra are analyzed separately. Age associations for the slow-timescale component (left) and fast-timescale component (right), shown separately for participants whose spectra were classified as one-knee or two-knee models. Each dot represents one participant, grouped by age range. Lines show the best-fitting model for each subset. For the slow-timescale component, both one-knee and two-knee spectra were best described by constant models, indicating no detectable age-related shift. By contrast, for the fast-timescale component, both one-knee and two-knee spectra were best described by linear models, indicating an age-related increase in fast knee frequency. These results show that the dissociation observed in the main analysis does not depend on whether the component was recovered from a one-knee or two-knee spectrum.
